# Reliability of self-reported risk factors for age-related brain disease

**DOI:** 10.1101/2024.04.25.24306370

**Authors:** Reinier W.P. Tack, Jasper R. Senff, Tamara N. Kimball, Savvina Prapiadou, Nirupama Yechoor, Jonathan Rosand, Sanjula D. Singh, Christopher D. Anderson

## Abstract

**Importance:** modifiable risk factors, including hypertension, hypercholesterolemia, diabetes, kidney disease, hearing impairment, and being overweight, increase risk of stroke, dementia, and late life depression (LLD), but may be poorly understood by patients.

**Objective:** To aid in the design of brain health risk stratification tools, we aimed to determine whether self-reported modifiable risk factors can identify at-risk patients on a population level.

**Design:** cohort study using cross-sectional data of ten iterations of the National Health and Examination Surveys (NHANES) from 1999 to 2018. Analyses were conducted between February and April of 2024.

**Setting:** US population-based cohorts

**Participants:** all participants who had both questionnaire data and objective measurements available.

**Exposure:** we collected data on answers to common questions and compared these to objectively measured risk factors for age-related brain disease.

**Main outcomes and measures:** we compared answers to simple questions to objectively measured risk factors. We reported means and 95% confidence intervals of objective measurements, created confusion matrices to determine common metrics, and compared performance of individual questions. We defined a question reliable if performance, measured through F1 scores, was > 0.7, as moderately reliable if F1 score was 0.5-0.7 and as unreliable if F1 score was < 0.5.

**Results:** Participants with both objective measurements and questionnaire data ranged from 16,966 (median age 32, 9,113 [20%] black, 19,993 [45%] white) in hearing impairment to 63,834 (median age 39, 14,156 [22%] black, 25,754 [40%] white) in diabetes. Mean values of objective measurements were significantly increased across all risk factors in patients that responded “Yes” to common questions on their presence compared to those who answered “No”. Performance of questions in identifying at-risk patients measured by F1 score was 0.25 in kidney disease, 0.44 in hypercholesterolemia, 0.56 in hypertension, 0.59 in hearing impairment, 0.71 in diabetes and 0.81 in being overweight.

**Conclusion and relevance:** in this study of multiple cohorts of up to 63,834 Americans, self-reported awareness of meeting clinical criteria for diabetes or being overweight were reliable, hypertension or hearing impairment were moderately reliable, and hypercholesterolemia or kidney disease were unreliable. These results could be used to guide construction of risk factor screening tools.

## Introduction

Age-related brain disease, including stroke, dementia and late-life depression (LLD), are a major cause of disability-adjusted life years (DALYs) ^1^. Modifiable risk factors have been identified as a key driver of the pathogenesis of these diseases. Recent estimates show that modifiable risk factors account for up to 40% of dementia incidence, 55% of LLD incidence and up to 90% of stroke incidence ^2–4^. As a result, advocacy organizations place these modifiable risk factors at the center of their platform, advocating for identification of at-risk individuals, education on modifiable risk factors, and direct-to-public messaging^5,6^.

Currently, modifiable risk factors are combined into predictive models, with the goal of identifying people at-risk for age-related brain disease^7–9^. These models often include hypertension^10–12^, hypercholesterolemia^13–15^, diabetes^16,17^, kidney disease^18,19^, being overweight^20,21^, and hearing impairment^22–24^. Previous studies have shown that people are often not aware of the presence of risk factors: only a small proportion of people was able to accurately recall their HbA1c or cholesterol levels (25% and 19% respectively)^25,26^. For blood pressure, erroneous reporting is common and the ability to accurately measure ones blood pressure at home remains limited^27,28^. These knowledge gaps limit the out-of-clinic applicability of models that include these risk factors and undermine the direct-to-public messaging goals of advocacy organizations aiming to improve population brain health.

Inclusion of self-reported risk factors could be one approach to democratize stratification tools while preserving model accuracy. However, it is currently unknown whether these self-reported risk factors can accurately identify people at-risk for age-related brain disease, and which manner of framing risk factor-related questions achieves the best performance in capturing these risk factors. Therefore, we aimed to determine the performance of self-reported risk factors for age-related brain disease in a representative US population sample, and which questions are most reliable in capturing risk information.

## Methods

### Data collection

The National Health and Nutrition Examination Survey (NHANES) is iteratively conducted by the National Center for Health and Statistics^29^. It consists of multiple cross-sectional data collections, where the design is focused on the representativeness of the cohort to the US population. Data collection methods include physical examinations, laboratory measurements, and at-home interviews. These data are publicly available and released in 2-year cycles. In this study, we analyzed data from ten iterations, between 1999 and 2018. We collected data on six self-reported risk factors, including demographics, responses to common questionnaires, and corresponding objective measurements.

### In and exclusion criteria

For analyses of individual risk factors, we included all participants where answers to common questions and objective measurements were available. Participants were excluded if they answered “Don’t know” or “Refused” to questions, or if data was missing. For hypertension analyses, we included participants where at least one blood pressure (both systolic and diastolic) measurement was available. For hearing impairment, we included participants where at least one measurement per frequency was available on audiogram.

### Subjective measurements

We included only questions and measurements that were consistent across iterations. For hypertension and diabetes questionnaire items following check items, where a follow-up question is only asked when participant respond “Yes” to the previous question, answers of non-responders for the second question were recategorized as “No”. For example, the question “Has a doctor ever told you more than once you had hypertension” is only asked to those who responded “Yes” to the question “Has a doctor ever told you you had hypertension?”. Participants who responded “No” to the initial question were recategorized as responding “No” to the follow-up question. For questions with multiple outcomes, such as “Would you say your hearing is excellent, good, that you have a little trouble, moderate trouble, a lot of trouble or are you deaf?”, answers were dichotomized at different cut-offs to evaluate performance (such as excellent vs all other answers). Where appropriate, answers to multiple questions were combined to evaluate performance of a combination of questions in predicting objective risk factors.

### Objective measurements

Each participant underwent one to three blood pressure readings, which were then averaged to obtain a more precise measurement. Hypertension was defined as a Systolic Blood Pressure (SBP) > 130 mmHg or a Diastolic Blood Pressure (DBP) of > 80 mmHg. Hypercholesterolemia was defined as a Total Cholesterol (TC) > 240 mg/dL. Diabetes was defined as HbA1c > 6.4%. Kidney disease was defined as a Glomerular Filtration Rate (GFR) < 60 ml/min/1.73m^2^ (estimated by MDRD-4^30^). Being overweight was defined as a Body Mass Index (BMI) > 25, calculated as weight (kg)/height(m)^2^. Hearing impairment was defined as a maximum decibel (dB) loss > 40 in speech frequencies.

### Statistical Analyses

We calculated mean, standard deviation (SD) and 95% confidence intervals (CI) using standard error (SE) of objective measurements, stratified by answers to common questions, and compared these through independent T-tests. Sensitivity analyses were performed to test for the robustness of our results using the interview standard sampling weights provided by the NHANES to adjust for non-response and post-stratification^31^. Confusion matrices were used to evaluate metrics (accuracy, sensitivity, specificity, positive predictive value (PPV), negative predictive value (NPV)) of self-reported risk factors in predicting dichotomized objective measurements. Performance of different questions was determined by comparison of F1-scores, calculated as previously described^32^. F1-scores are a harmonic mean of precision (sensitivity) and recall (PPV), and are measures of a question’s ability to accurately identify patients where an objective risk factor is present. We defined poor performance as an F1-score < 0.5, moderate performance as an F1-score 0.5-0.7 and good performance as an F1-score > 0.7^33^. As a sensitivity analysis, we plotted F1 score against different cut-off values for our included risk factors in order to visualize the cut-off value of the risk factor for which the questions optimally dichotomized the sample. All analyses were performed using R studio, version 4.1.0.

### Ethical approval

The NHANES has previously obtained research ethics board approval. Written informed consent has been obtained for all subjects included in the study.

## Results

### Hypertension

Demographic characteristics of participants are shown in Supplementary table 1. In total, 57,433 participants had at least one blood pressure measurement and questionnaire data, and 17,601 participants responded “Yes” to at least one question (Supplementary table 2). We excluded 104 participants who answered “Don’t Know”. Mean SBP ranged from 135 (95% CI 134.9 – 135.1) to 136 (95% CI 135.6 – 136.3) when answering “Yes” to common questions and from 118 (95% CI 117.9 - 118.1) to 120 (95% CI 119.9-120.1) when answering “No”. Mean DBP ranged from 70.0 (95% CI 69.6 – 70.4) to 71.4 (95% CI 71.0 – 71.8) when answering “Yes” to common questions and from 68.1 (95% CI 67.9 – 68.1) to 68.8 (95% CI 68.6 – 67.0) when answering “No” (Figure 1). Sensitivity analysis using weighted means did not alter the results (Supplementary table 3). Performance of different questions was moderate; F1 score ranged from 0.48 to 0.56 (Figure 2). Sensitivity analyses using different cut-offs showed that optimal performance was achieved at an SBP between 120-125 and a DBP >80 (Supplementary Figure 1).

**Figure 1.**
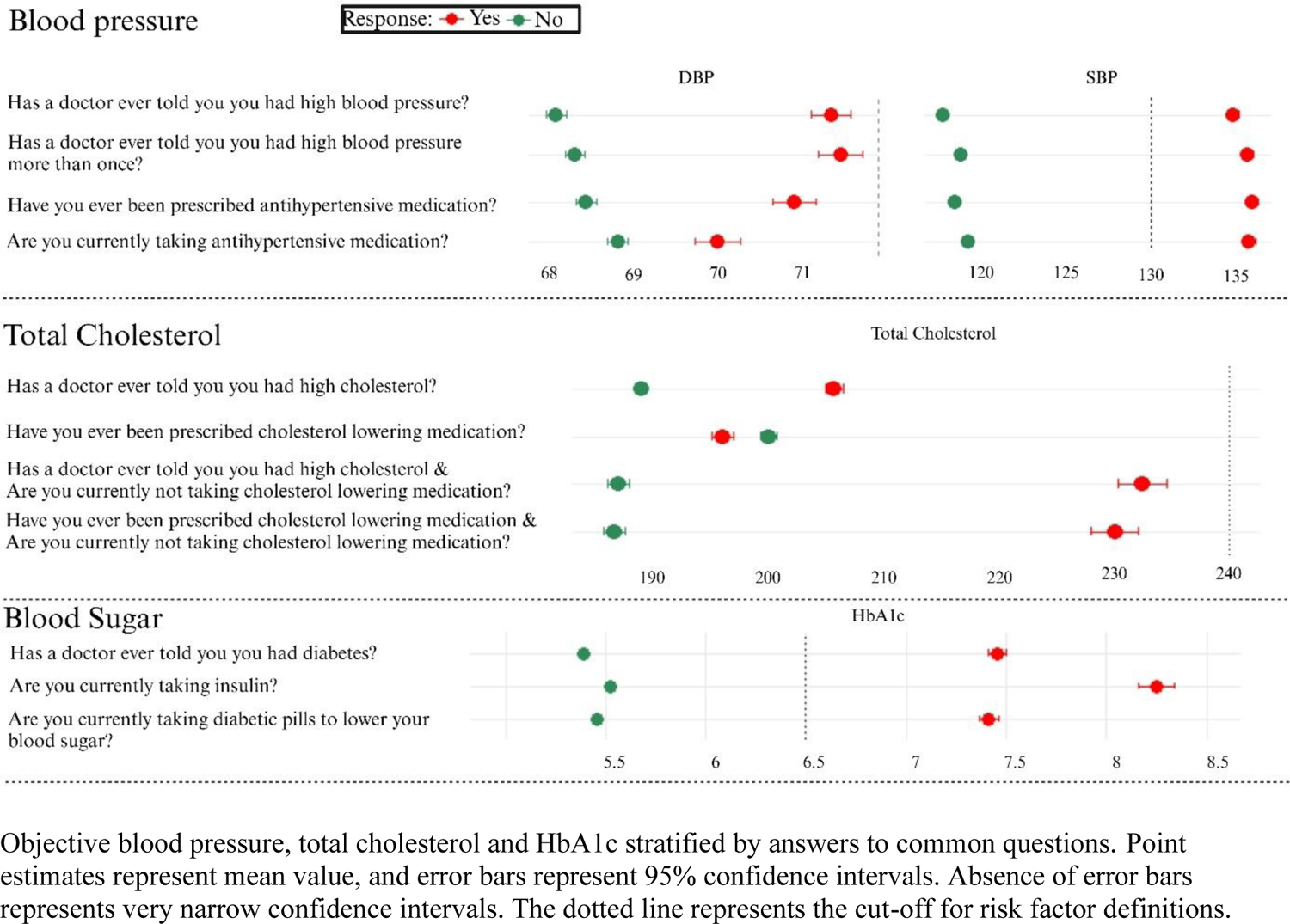
Blood pressure, Cholesterol and HbA1c stratified by answers to common questions.

**Figure 2.**
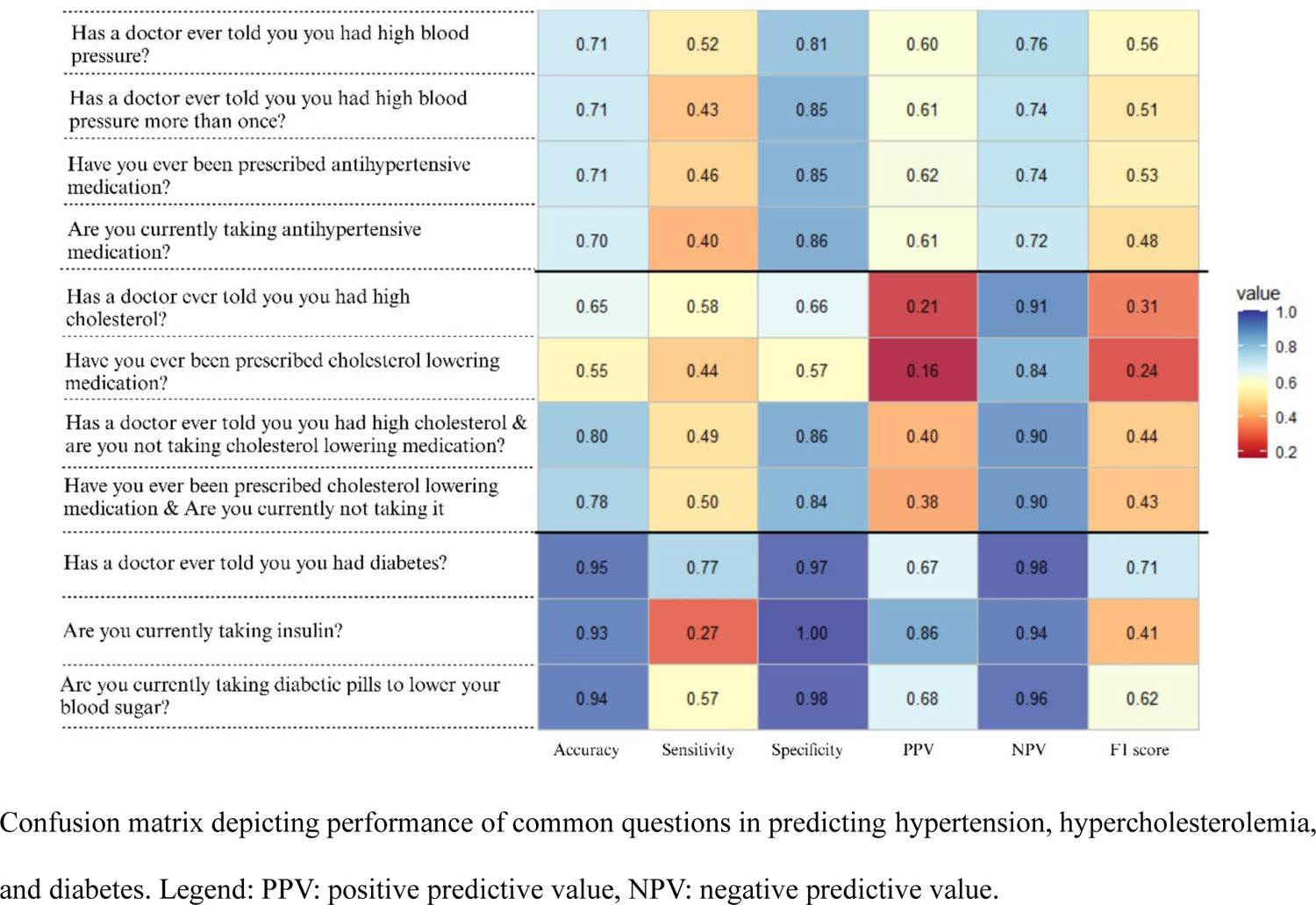
Performance of questions for hypertension, hypercholesterolemia, and diabetes

### Hypercholesterolemia

Demographic characteristics of participants are shown in Supplementary table 1. In total, 41,535 participants had cholesterol measurement and questionnaire data, and 15,400 participants responded “Yes” to at least one question (Supplementary table 2). We excluded 294 patients who answered “Don’t Know” or refused to answer. Mean TC ranged from 196 (95% CI 195-197) to 232 (95% CI 230 - 235) when answering “Yes” to one or a combination of questions and from 186 (95% CI 185 - 187) to 200 (95% CI 199 - 201) when answering “No” (Figure 1). Sensitivity analysis using weighted means did not alter the results (Supplementary table 3). Performance of individual questions was poor (F1 score 0.24 to 0.31), but improved somewhat when combining the question “Has a doctor ever told you you have high cholesterol?” with the question “Are you currently taking cholesterol lowering medication?” (F1 score 0.44) (Figure 2). Sensitivity analyses using different cutoffs showed that performance of individual questions was far better at lower cut-off values (between 100-150), and performance of a combined question was optimal between 200 and 225 (Supplementary Figure 2). A second sensitivity analysis showed that using Low-Density Lipoprotein (LDL) as a measure of hypercholesterolemia resulted in worse performance (F1 score < 0.3) (Supplementary Figure 2).

### Diabetes

Demographic characteristics of participants are shown in Supplementary table 1. In total, 63,834 participants had an HbA1c measurement and questionnaire data, and 6,124 participants responded “Yes” to at least one question (Supplementary table 2). We excluded 57 participants who answered “Don’t Know”, refused to answer or answered borderline. Mean HbA1c ranged from 7.41 (95% CI 7.32 – 7.50) to 8.25 (95% CI 8.11 – 8.39) when answering “Yes” to common questions and from 5.39 (95% CI 5.38 – 5.40) to 5.52 (95% CI 5.51 – 5.53) when answering “No” (Figure 1). Sensitivity analysis using weighted means did not alter the results (Supplementary table 3). Performance was good when asking the question “Has a doctor ever told you you have Diabetes?” (F1 score 0.71) (Figure 2). Sensitivity analysis using different cutoffs showed that performance is optimal at a HbA1c between 6.2 and 6.4 (Supplementary Figure 3).

### Kidney Disease

Demographic characteristics of participants are shown in Supplementary table 4. In total, 44,813 participants had questionnaire data and sufficient data to calculate GFR, and 2,962 participants responded “Yes” to at least one question (Supplementary table 5). We excluded 98 participants who answered “Don’t Know” or refused to answer. Mean GFR ranged from 61.1 (95% CI 59.1 – 63.1) to 83.6 (95% CI 83.3 – 83.9) when answering “Yes” to common questions and from 90.2 (95% CI 89.9 – 90.5) to 91.1 (95% CI 90.8 – 91.4) when answering “No” (Figure 3). Sensitivity analysis using weighted means did not alter the results (Supplementary table 6). Performance of individual questions was poor (F1 score 0.16-0.25) (Figure 4). Sensitivity analysis showed that performance was optimal at a GFR of 40-45 (Supplementary Figure 4).

**Figure 3.**
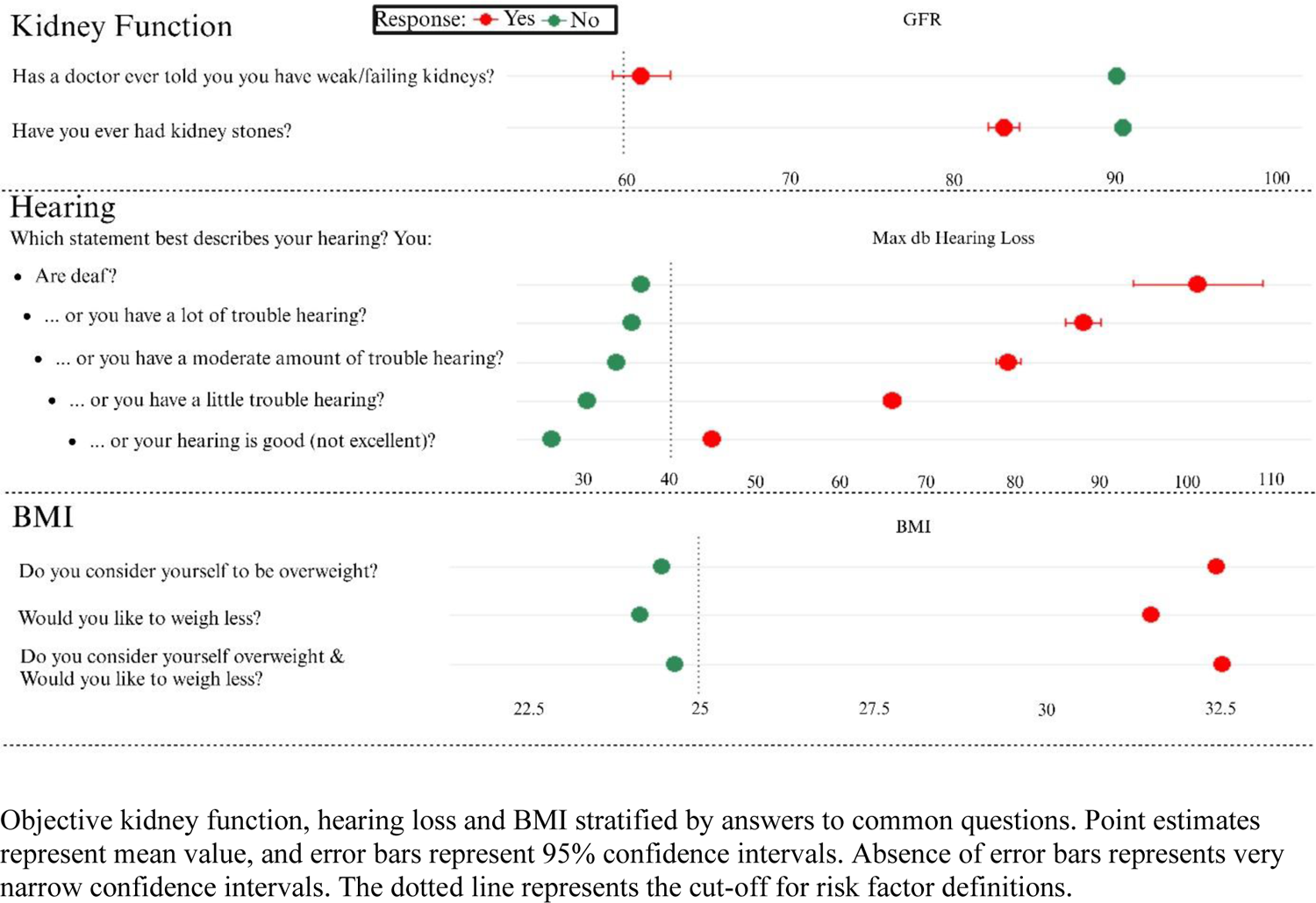
GFR, hearing loss, and BMI stratified by answers to common questions.

**Figure 4.**
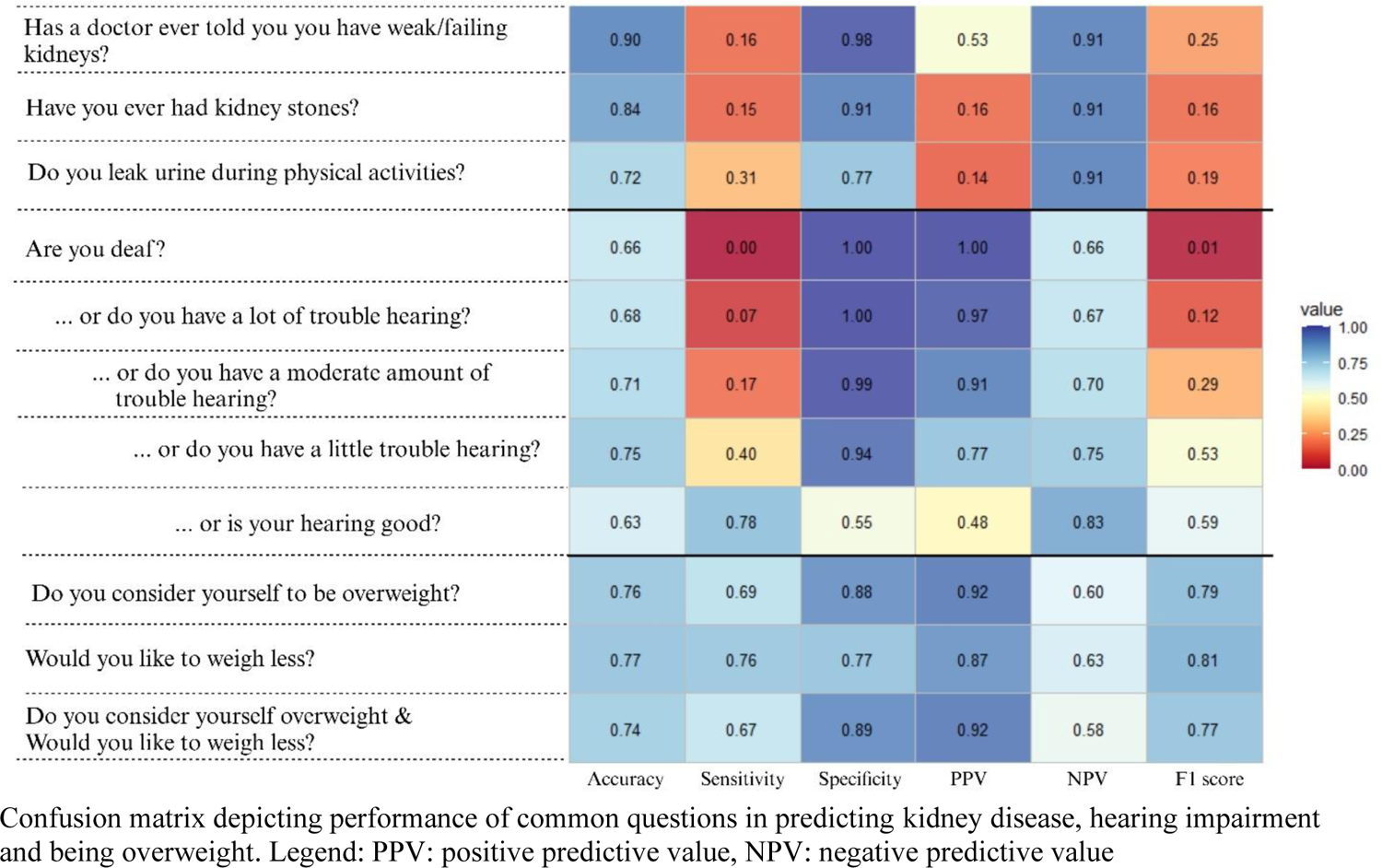
Performance of questions for kidney disease, hearing impairment, and being overweight

### Hearing impairment

Demographic characteristics of participants are shown in Supplementary table 4. In total, 16,966 participants had an audiogram performed and questionnaire data (Supplementary table 5). We included only the question: “Would you say your hearing is excellent, good, that you have a little trouble, moderate trouble, a lot of trouble or are you deaf?” and compared mean of maximum dB loss across different cut-offs (deaf vs all others, deaf or a lot of trouble vs all others, etc). We excluded 21 participants who answered “Don’t Know” or refused to answer. Hearing loss was consistent across different answers and was most pronounced at higher frequencies (Supplementary Figure 5). Hearing loss ranged from 101.0 dB (95% CI 95.1 – 106.9) to 44.8 (95% CI 44.0 – 45.6) across cut-offs (Figure 3). Sensitivity analysis using weighted means did not alter the results (Supplementary table 6).

Performance improved as cut-offs were adjusted, where the best performance was achieved when comparing participants that answered “Excellent” to all other answers (F1 score 0.59) (Figure 4).

Sensitivity analysis using different cutoffs showed that performance, when comparing those with “Excellent” hearing vs all other answers, was highest at low frequencies. When comparing those that answered “Excellent” or “Good” vs all others, performance was highest at a dB loss between 45-65 (Supplementary Figure 6).

### Overweight

Demographic characteristics of participants are shown in Supplementary table 4. In total, 59,319 participants had height and weight measurements as well as questionnaire data. We included the questions “Do you consider yourself to be overweight, underweight or about the right weight?” and “Would you like to weigh more, less or the same?” and dichotomized based on answering “Overweight” vs all other responses and “Less” vs all other responses (Supplementary table 5). We excluded 177 participants who answered “Don’t Know” or refused to answer. Mean BMI ranged from 31.5 (95% CI 31.4 – 31.6) to 32.6 (95% CI 32.5 – 32.7) when answering “Less” or “Overweight” and from 24.1 (95% CI 24.1-24.2) to 24.7 (95% CI 24.7-24.8) when answering “The same”, “More”, “Normal weight” or “Underweight” (Figure 3). Sensitivity analysis using weighted means did not alter the results (Supplementary table 6). Performance of individual questions was good (F1 score 0.77 – 0.81) (Figure 4). Sensitivity analysis using different cutoffs showed that performance was highest when using a BMI cutoff value between 24 and 26 (Supplementary Figure 7).

### Best performing questions

Table 1 shows the questions that performed best in assessment of modifiable risk factors based on F1-score.

**Table 1.** Best performing questions in assessing modifiable risk factors.

| **Risk factor** | **Recommended question** | **Performance** |
| --- | --- | --- |
| Hypertension | Has a doctor ever told you you had high blood pressure? | Moderate |
| Hypercholesterolemia | Has a doctor ever told you you had high cholesterol & are you not taking cholesterol lowering medication? | Poor |
| Diabetes | Has a doctor ever told you you had diabetes? | Good |
| Kidney disease | Has a doctor ever told you you have weak/failing kidneys? | Poor |
| Hearing impairment | Would you describe your hearing as excellent? | Moderate |
| Being overweight | Would you like to weigh less? | Good |

## Discussion

Our results show that assessment of self-reported modifiable risk factors for age-related brain disease through common questions leads to a significant dichotomization at the population level, but performance of these questions in identifying at-risk patients is remarkably variable between risk factors. Overall, performance of self-reported risk factor assessment was poor in assessing hypercholesterolemia and kidney disease, moderate in assessing hypertension and hearing impairment and good in assessing diabetes and being overweight. While the purpose of our study was to aid in the development of tools targeting risk factors for stroke, dementia, and LLD, there is substantial overlap between the studied risk factors and other common diseases such as coronary artery disease. As such, our results could inform on development of screening tools for other conditions as well.

Previous studies have reported similar findings in other populations. A meta-analysis of self-reported risk factors in a general population showed that the sensitivity was similar in assessment of hypertension (0.40-0.78), hypercholesterolemia (0.41-0.51), diabetes (0.68 – 0.80) and being overweight (0.72-0.84)^34^. Furthermore, in a previous study using data from the national registry cohort of 64,961 women, agreement between self-reported and measured risk factors (assessed through Cohen’s Kappa) was 0.71 for diabetes and 0.46 for hypertension, similar to our study, but 0.14 in hypercholesterolemia, far lower than our reported performance^35^. This difference might be caused by the increased performance achieved by combining multiple questions but might also be caused by the fact that objective measurements were previously obtained through documentation in hospital medical records, where under-recording of risk factors is well-known^36^.

We found that optimal performance of included risk factors and questions was achieved when using an SBP of 120-125 mmHg, a TC of 200-225 mg/dL, an HbA1c of 6.2-6.4%, and a BMI of 24-26 as cut-off values. This suggests these questions perform best when dichotomizing around a cut-off that includes patients with limited to moderate severity of risk factors, such as prehypertension or prediabetes. As such, these self-reported questions appear to be most accurately identifying patients who may be on the threshold of developing a risk factor for age-related brain disease, and may thereby serve as an effective tool for identifying individuals who might benefit from early clinical interventions.

A recent study on hearing impairment, using a definition as having either poor or fair hearing on a 5-point Likert scale or having moderate or great difficulty following a conversation if there is background noise, reported on a sensitivity of 0.79 when using the combination of these questions^37^.

Our data shows that a similar performance (0.78) can be achieved by asking a single question and dichotomizing by patients defining their hearing as “Excellent” or any other answer.

We found a low performance of questions assessing kidney disease. A previous study using general practitioner data in older adults also showed an agreement (Cohen’s Kappa) of 0.15 between self-reported and measured kidney disease^38^. Similarly, previous studies using known chronic kidney disease (CKD) patients report an awareness of kidney disease between 40-60%, which increased as their GFR decreased to <45^39,40^. This is in line with our results, which show that the performance of the questions is highest when using a GFR cutoff value between 35 and 45, and highlighting the fact that patients are generally not aware of their kidney disease before reaching stage 3B kidney disease. Our results and the previous literature show that self-reported kidney disease is insufficient in identifying patients at-risk, especially given the importance of early identification and treatment of chronic kidney disease^41^. Future research should focus on identifying questions with increased sensitivity for identifying kidney disease and an increased PPV for hypercholesterolemia, in addition to efforts to better educate patients on the importance of awareness and active management of these conditions.

Our study has limitations. First, we included only a limited number of questions, many of which included “Has a doctor…” terms, which might have caused the limited sensitivity in some questions. This is due to our being limited to the structure of questions included in NHANES materials. Second, while we show that some self-reported risk factors show good performance, we cannot make assumptions about the validity given the cross-sectional design of the NHANES dataset. Ideally, we would prospectively compare the impact of each self-reported risk factor on age-related brain disease to the impact of objective measurements. The strengths of our study are the size and representativeness of the included population, the novelty of comparing different questions head-to-head, and the use of F1 scores as a performance metric. We contend that F1 scores are more suited to assess the performance of self-reported risk factors than Cohen’s Kappa, as the relative cost of false negatives outweighs the benefit of correctly classifying true negative cases.

## Conclusion

Self-reported diabetes or being overweight is reliable, self-reported hypertension and hearing impairment is moderately reliable and self-reported hypercholesterolemia and kidney disease is unreliable. Future research should focus on asking the right questions to identify at-risk patients and assess the relative performance of models incorporating self-reported risk factors to conventional medical terminology in stratifying risk.

## Data Availability

All data produced in the present work are contained in the manuscript

