## Supplementary Material for "Reliability of self-reported risk factors for age-related brain disease"

**Supplementary Materials** **Page**

Supplementary Tables:

Supplementary Table 1. 2

Supplementary Table 2. 3

Supplementary Table 3. 4

Supplementary Table 4. 5

Supplementary Table 5. 6

Supplementary Table 6. 7

Supplementary Figures:

Supplementary Figure 1. 8

Supplementary Figure 2. 9

Supplementary Figure 3. 10

Supplementary Figure 4. 11

Supplementary Figure 5. 12

Supplementary Figure 6. 13

Supplementary Figure 7. 14

**Supplementary Table 1.**

| Question | Age | Female | Race | | | | |
|---|---|---|---|---|---|---|---|
| | | | Mexican American | Other Hispanic | White | Black | Other |
| **Blood Pressure** | | | | | | | |
| Has a doctor ever told you you had high blood pressure? | 44 (16-85) | 29,465 (51%) | 10,821 (19%) | 4,594 (8%) | 24,166 (42%) | 12,646 (22%) | 5,225 (9%) |
| Has a doctor ever told you >1 you had high blood pressure? | 44 (16-85) | 29,434 (51%) | 10,809 (19%) | 4,592 (8%) | 24,127 (42%) | 12,632 (22%) | 5,217 (9%) |
| Have you ever been prescribed antihypertensive medication? | 44 (16-85) | 29,457 (51%) | 10,815 (19%) | 4,594 (8%) | 24,159 (42%) | 12,644 (22%) | 5,221 (9%) |
| Are you currently taking antihypertensive medication? | 44 (16-85) | 29,456 (51%) | 10,815 (19%0 | 4,594 (8%) | 24,155 (42%) | 12,644 (22%) | 5,221 (9%) |
| **Cholesterol** | | | | | | | |
| Has a doctor ever told you you had high cholesterol? | 51 (16-85) | 21,776 (52%) | 6,303 (15%) | 3,504 (8%) | 18,922 (46%) | 8,428 (20%) | 4,378 (11%) |
| Have you ever been prescribed cholesterol lowering medication? | 56 (16-85) | 12,791 (52%) | 3,335 (14%) | 2,181 (9%) | 10,924 (45%) | 4,997 (21%) | 2,938 (12%) |
| Has a doctor ever told you you had high cholesterol & Are you currently not taking cholesterol lowering medication? | 64 (16-85) | 5,109 (49%) | 1,477 (14%) | 940 (9%) | 5201 (49%) | 1,973 (19%) | 922 (9%) |
| Have you ever been prescribed cholesterol-lowering meds & Are you not currently taking it? | 64 (16-85) | 5,109 (49%) | 1,477 (14%) | 940 (9%) | 5201 (49%) | 1,973 (19%) | 922 (9%) |
| **Blood Sugar** | | | | | | | |
| Has a doctor ever told you you had diabetes? | 39 (12-85) | 32,714 (51%) | 13,119 (21%) | 5,097 (8%) | 25,754 (40%) | 14,156 (22%) | 5,708 (9%) |
| Are you currently taking insulin? | 40 (12-85) | 32,711 (51%) | 13,118 (21%) | 5,097 (8%) | 25,750 (40%) | 14,155 (22%) | 5,708 (9%) |
| Are you currently taking diabetic pills to lower your blood sugar? | 39 (12-85) | 32,706 (51%) | 13,113 (21%) | 5,096 (8%) | 25,748 (40%) | 14,153 (22%) | 5,707 (9%) |

Demographics of included participants. Age is shown as median (range), other variables as number (percentage). "Other" race includes multi-racial.

**Supplementary table 2.**

| **Blood pressure** | | | | | | | |
|---|---|---|---|---|---|---|---|
| Question | N (total) | N (Yes) | N (No) | Mean SBP "Yes" (SD) | Mean SBP "No" (SD) | Mean DBP "Yes" (SD) | Mean DBP "No" (SD) |
| Has a doctor ever told you you had high blood pressure? | 57452 | 17601 | 39851 | 135 (21.3)* | 118 (15.8) | 71.3 (15.7)* | 68.1 (12.2) |
| Has a doctor ever told you >1 you had high blood pressure? | 57377 | 14201 | 43176 | 136 (21.6)* | 119 (16.5) | 71.4 (15.8)* | 68.3 (12.5) |
| Have you ever been prescribed antihypertensive medication? | 57433 | 14927 | 42506 | 136 (21.5)* | 119 (16.2) | 70.9 (16.0)* | 68.4 (12.4) |
| Are you currently taking antihypertensive medication? | 57429 | 13025 | 44404 | 136 (21.4)* | 120 (17.0) | 70.0 (15.8) † | 68.8 (12.7) |

| **Cholesterol** | | | | | |
|---|---|---|---|---|---|
| Question | N (total) | N (Yes) | N (No) | Mean TC "Yes" (SD) | Mean TC "No" (SD) |
| Has a doctor ever told you you had high cholesterol? | 41535 | 15400 | 26135 | 205 (47.5)* | 188 (38.9) |
| Have you ever been prescribed cholesterol lowering medication? | 24375 | 10526 | 13849 | 196 (48.9) † | 200 (41.9) |
| Has a doctor ever told you you had high cholesterol & Are you currently not taking cholesterol lowering medication? | 10513 | 2110 | 8403 | 232 (50.7)* | 186 (43.9) |
| Have you ever been prescribed cholesterol-lowering meds & Are you not currently taking it? | 10513 | 2287 | 8226 | 230 (50.5)* | 186 (43.9) |

| **Blood Sugar** | | | | | |
|---|---|---|---|---|---|
| Question | N (total) | N (Yes) | N (No) | Mean HbA1c "Yes" (SD) | Mean HbA1c "No" (SD) |
| Has a doctor ever told you you had diabetes? | 63834 | 6124 | 57710 | 7.46 (1.83)* | 5.39 (0.57) |
| Are you currently taking insulin? | 63158 | 1722 | 62106 | 8.25 (1.91)* | 5.52 (0.85) |
| Are you currently taking diabetic pills to lower your blood sugar? | 63817 | 4497 | 59320 | 7.41 (1.69)* | 5.46 (0.75) |

Differences in blood pressure, cholesterol and blood sugar measurements between participants answering "Yes" or "No" for specific questions for blood pressure. Significance level from independent t-test are shown (*$p < 5 \times 10^{-16}$, †$p < 5 \times 10^{-6}$, \`$p < 5 \times 10^{-4}$). Legend: SBP: Systolic Blood Pressure; DBP: Diastolic Blood Pressure; TC: Total Cholesterol; HbA1c: Hemoglobin A1c; SD: Standard Deviation.

**Supplementary table 3.**

| Blood pressure (weighted) | | | | | | | |
|---|---|---|---|---|---|---|---|
| Question | N (total) | N (Yes) | N (No) | Mean SBP "Yes" (SD) | Mean SBP "No" (SD) | Mean DBP "Yes" (SD) | Mean DBP "No" (SD) |
| Has a doctor ever told you you had high blood pressure? | 57452 | 17601 | 39851 | 135 (21.3)* | 118 (15.8) | 72.4 (14.7)* | 69.6 (11.4) |
| Has a doctor ever told you >1 you had high blood pressure? | 57377 | 14201 | 43176 | 133 (20.1)* | 118 (15.3) | 72.5 (14.9)* | 69.7 (11.6) |
| Have you ever been prescribed antihypertensive medication? | 57433 | 14927 | 42506 | 133 (20.3)* | 118 (15.1) | 71.9 (15.0)* | 69.9 (11.5) |
| Are you currently taking antihypertensive medication? | 57429 | 13025 | 44404 | 133 (20.2)* | 119 (15.7) | 71.0 (14.9)` | 70.2 (11.8) |

| Cholesterol (weighted) | | | | | |
|---|---|---|---|---|---|
| Question | N (total) | N (Yes) | N (No) | Mean TC "Yes" (SD) | Mean TC "No" (SD) |
| Has a doctor ever told you you had high cholesterol? | 41535 | 15400 | 26135 | 207.9 (47.5)* | 189.4 (37.7) |
| Have you ever been prescribed cholesterol lowering medication? | 24375 | 10526 | 13849 | 197.1 (49.5) † | 201.7 (41.5) |
| Has a doctor ever told you you had high cholesterol & Are you currently not taking cholesterol lowering medication? | 10513 | 2110 | 8403 | 235.6 (51.5)* | 186.8 (43.5) |
| Have you ever been prescribed cholesterol-lowering meds & Are you not currently taking it? | 10513 | 2287 | 8226 | 233.7 (51.2)* | 186.4 (43.4) |

| Blood Sugar (Weighted) | | | | | |
|---|---|---|---|---|---|
| Question | N (total) | N (Yes) | N (No) | Mean HbA1c "Yes" (SD) | Mean HbA1c "No" (SD) |
| Has a doctor ever told you you had diabetes? | 63834 | 6124 | 57710 | 7.35 (1.74)* | 5.36 (0.52) |
| Are you currently taking insulin? | 63158 | 1722 | 62106 | 8.16 (1.83)* | 5.47 (0.75) |
| Are you currently taking diabetic pills to lower your blood sugar? | 63817 | 4497 | 59320 | 7.30 (1.60)* | 5.41 (0.68) |

Weighted differences in blood pressure, cholesterol and blood sugar measurements between participants answering "Yes" or "No" for specific questions for blood pressure. Significance level from independent t-test are shown (*$p < 5 \times 10^{-16}$, †$p < 5 \times 10^{-6}$, \`$p < 5 \times 10^{-4}$). Legend: SBP: Systolic Blood Pressure; DBP: Diastolic Blood Pressure; TC: Total Cholesterol; HbA1c: Hemoglobin A1c; SD: Standard Deviation.

**Supplementary Table 4.**

| Question | Age | Female | Race | | | | |
|---|---|---|---|---|---|---|---|
| | | | Mexican American | Other Hispanic | White | Black | Other |
| **Kidney Function** | | | | | | | |
| Has a doctor ever told you you have weak/failing kidneys? | 49 (20-85) | 23,124 (52%) | 7,569 (17%) | 3,780 (8%) | 19,993 (45%) | 9,113 (20%) | 4,358 (10%) |
| Have you ever had kidney stones? | 50 (20-80) | 16,068 (51%) | 4,769 (15%) | 3,291 (11%) | 12,875 (41%) | 6,426 (21%) | 3,842 (12%) |
| **Hearing** | | | | | | | |
| Would you say your hearing is excellent, good, that you have a little trouble, moderate trouble, a lot of trouble or that you are deaf? | 32 (6-85) | 8,454 (50%) | 3,018 (18%) | 1,561 (9%) | 6,170 (36%) | 4,089 (24%) | 2,128 (13%) |
| **Body Mass Index** | | | | | | | |
| Do you consider yourself to be overweight? | 44 (16-85) | 30,495 (51%) | 11,244 (19%) | 4,756 (8%) | 24,656 (42%) | 13,110 (22%) | 5,475 (9%) |
| Would you like to weigh less? | 44 (16-85) | 30,536 (51%) | 11,290 (19%) | 4,772 (8%) | 24,658 (42%) | 13,115 (22%) | 5,484 (9%) |
| Do you consider yourself to be overweight & would you like to weigh less? | 44 (16-85) | 30,484 (51%) | 11,241 (19%) | 4,753 (8%) | 24,640 (42%) | 13,107 (22%) | 5,475 (9%) |

Demographics of included participants. Age is shown as median (range), other variables as number (percentage). “Other” race includes multi-racial.

**Supplementary table 5.**

| **Kidney Function** | | | | | |
|---|---|---|---|---|---|
| Question | N (total) | N (Yes) | N (No) | Mean GFR "Yes" (SD) | Mean GFR "No" (SD) |
| Has a doctor ever told you you have weak/failing kidneys? | 44813 | 1404 | 43409 | 61.1 (34.2)* | 90.6 (26.6) |
| Have you ever had kidney stones? | 31203 | 2962 | 28241 | 83.6 (26.4)* | 91.1 (26.4) |
| **Hearing** | | | | | |
| Question | N (total) | N (Yes) | N (No) | Mean db loss "Yes" (SD) | Mean db loss "No" (SD) |
| Are you deaf? | 16966 | 20 | 16946 | 101.0 (15.9)* | 36.6 (26.3) |
| … or do you have a lot of trouble hearing? | 16966 | 392 | 16574 | 87.7 (20.5)* | 35.4 (25.2) |
| … or do you have moderate moderate trouble hearing? | 16966 | 1111 | 15855 | 79.1 (24.8)* | 33.7 (23.7) |
| … or do you have a little trouble hearing? | 16966 | 3027 | 13939 | 65.6 (28.9)* | 30.4 (21.0) |
| … or is your hearing good (not excellent) | 16966 | 9545 | 7421 | 44.8 (28.9)* | 26.2 (17.9) |
| **Body Mass Index** | | | | | |
| Question | N (total) | N (Yes) | N (No) | Mean BMI "Yes" (SD) | Mean BMI "No" (SD) |
| Do you consider yourself to be overweight? | 59241 | 29341 | 29900 | 32.5 (6.7)* | 24.5 (4.2) |
| Would you like to weigh less? | 59319 | 34430 | 19475 | 31.5 (6.8)* | 24.1 (4.2) |
| Do you consider yourself to be overweight & would you like to weigh less? | 59216 | 28251 | 30965 | 32.6 (6.7)* | 24.7 (4.4) |

Differences in GFR, hearing impairment and BMI between participants answering "Yes" or "No" for specific questions. Significance level from independent t-test are shown (*$p < 5 \times 10^{-16}$, †$p < 5 \times 10^{-6}$, \`$p < 5 \times 10^{-4}$). Legend: GFR: Glomerular Filtration Rate; db: decibels; BMI: Body Mass Index; SD: Standard Deviation.

**Supplementary table 6.**

| **Kidney Function (Weighted)** | | | | | |
|---|---|---|---|---|---|
| Question | N (total) | N (Yes) | N (No) | Mean GFR "Yes" (SD) | Mean GFR "No" (SD) |
| Has a doctor ever told you you have weak/failing kidneys? | 44813 | 1404 | 43409 | 65.0 (32.8)* | 89.0 (25.0) |
| Have you ever had kidney stones? | 31203 | 2962 | 28241 | 83.7 (23.8)* | 90.0 (24.2) |
| **Hearing (Weighted)** | | | | | |
| Question | N (total) | N (Yes) | N (No) | Mean db loss "Yes" (SD) | Mean db loss "No" (SD) |
| Are you deaf? | 16966 | 20 | 16946 | 101.9 (15.1)* | 38.0 (25.6) |
| … or do you have a lot of trouble hearing? | 16966 | 392 | 16574 | 87.6 (20.3)* | 36.9 (24.6) |
| … or do you have moderate moderate trouble hearing? | 16966 | 1111 | 15855 | 77.8 (24.0)* | 35.1 (23.2) |
| … or do you have a little trouble hearing? | 16966 | 3027 | 13939 | 64.0 (27.6)* | 31.6 (20.6) |
| … or is your hearing good (not excellent) | 16966 | 9545 | 7421 | 45.6 (27.6)* | 27.3 (17.7) |
| **Body Mass Index (Weighted)** | | | | | |
| Question | N (total) | N (Yes) | N (No) | Mean BMI "Yes" (SD) | Mean BMI "No" (SD) |
| Do you consider yourself to be overweight? | 59241 | 29341 | 29900 | 32.1 (6.6)* | 24.2 (4.0) |
| Would you like to weigh less? | 59319 | 34430 | 19475 | 31.1 (6.7)* | 23.9 (4.0) |
| Do you consider yourself to be overweight & would you like to weigh less? | 59216 | 28251 | 30965 | 32.2 (6.6)* | 24.4 (4.2) |

Weighted differences in GFR, hearing impairment and BMI between participants answering "Yes" or "No" for specific questions. Significance level from independent t-test are shown (*$p < 5 \times 10^{-16}$, †$p < 5 \times 10^{-6}$, \`$p < 5 \times 10^{-4}$). Legend: GFR: Glomerular Filtration Rate; db: decibels; BMI: Body Mass Index; SD: Standard Deviation.

### Supplementary Figure 1

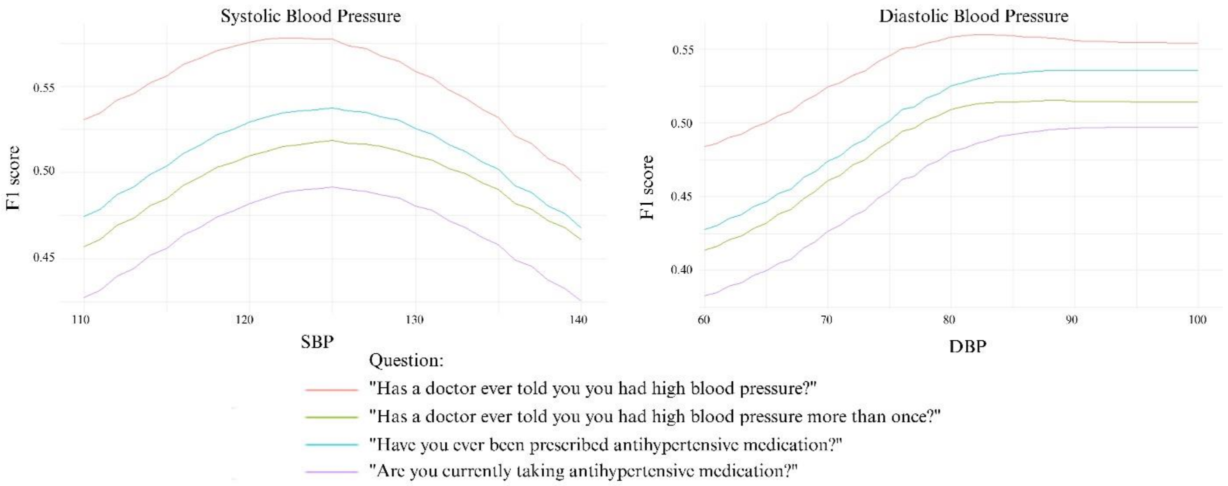


F1 score for different cut-off values of systolic and diastolic blood pressure for common questions.

**Supplementary Figure 2**

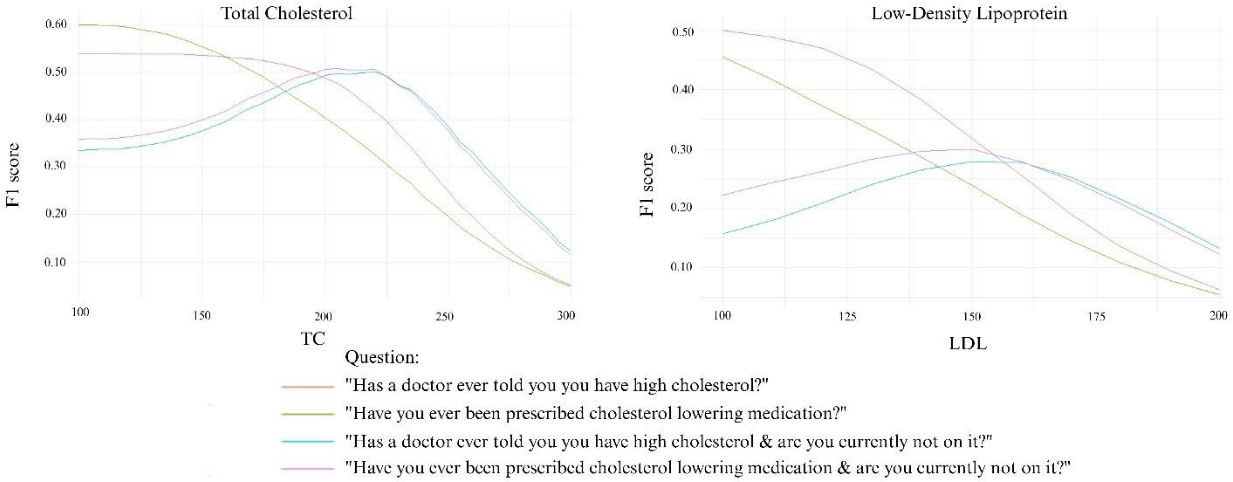


F1 score for different cut-off values of total cholesterol and LDL for common questions.

**Supplementary Figure 3**

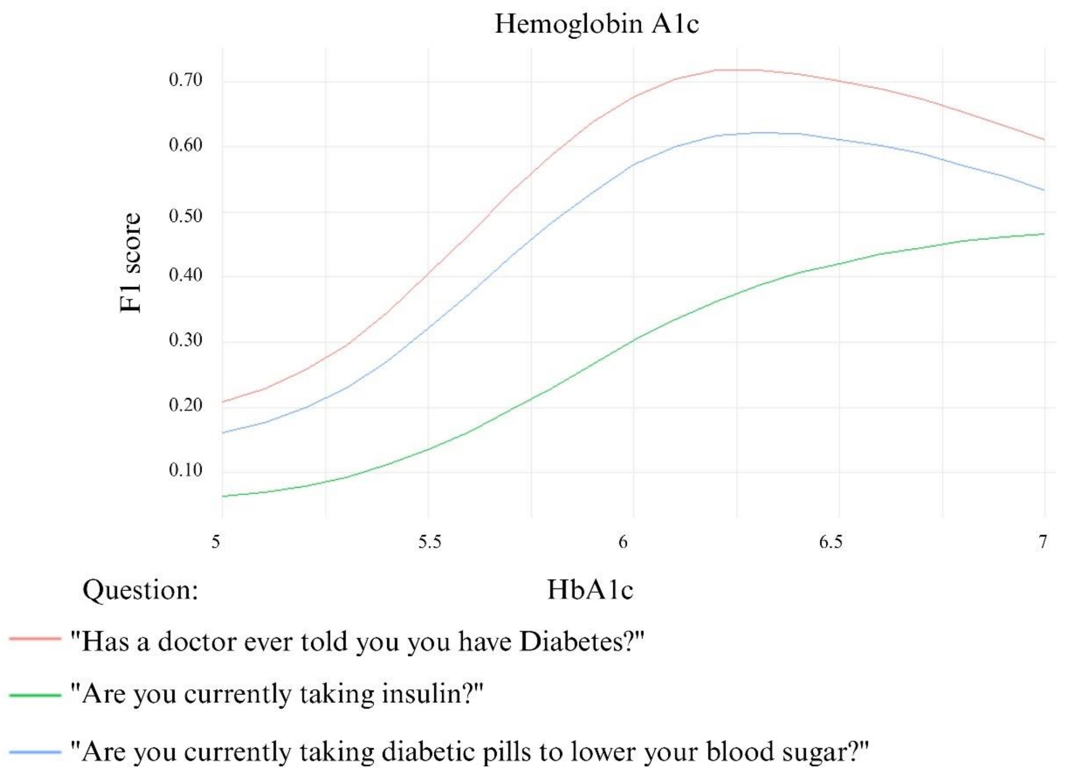


F1 score for different cut-off values of HbA1c for common questions.

**Supplementary Figure 4**

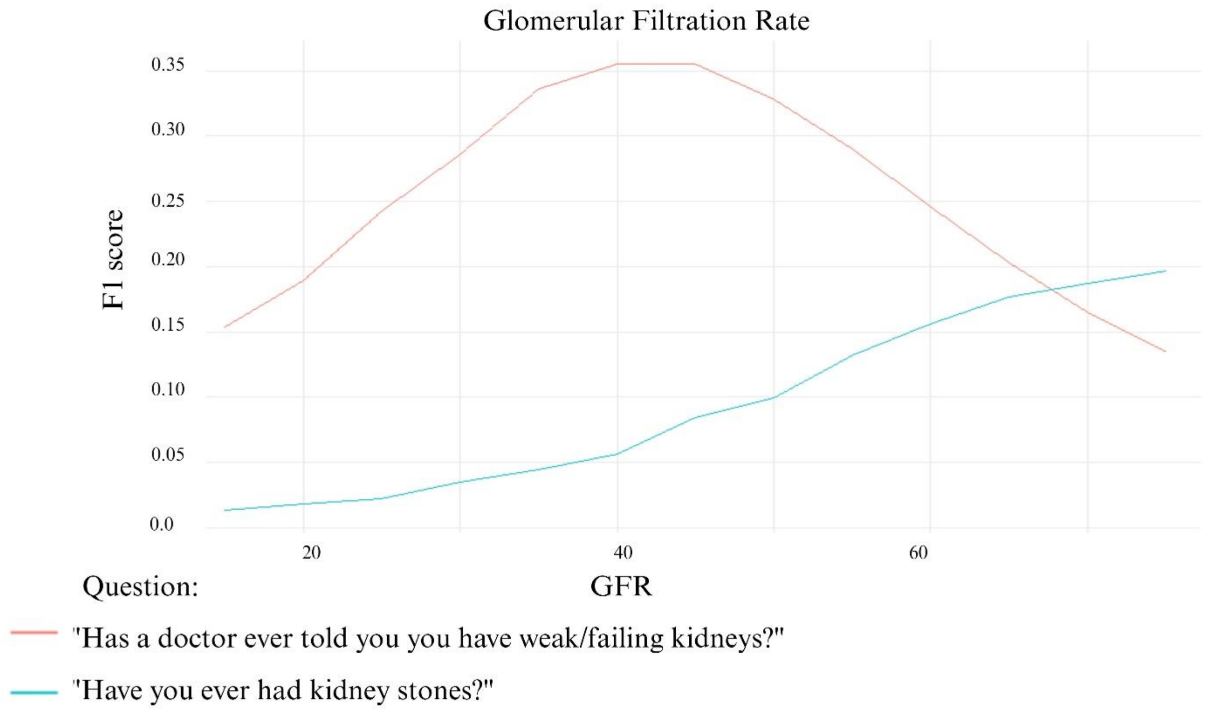


F1 score for different cut-off values of GFR for common questions.

**Supplementary Figure 5**

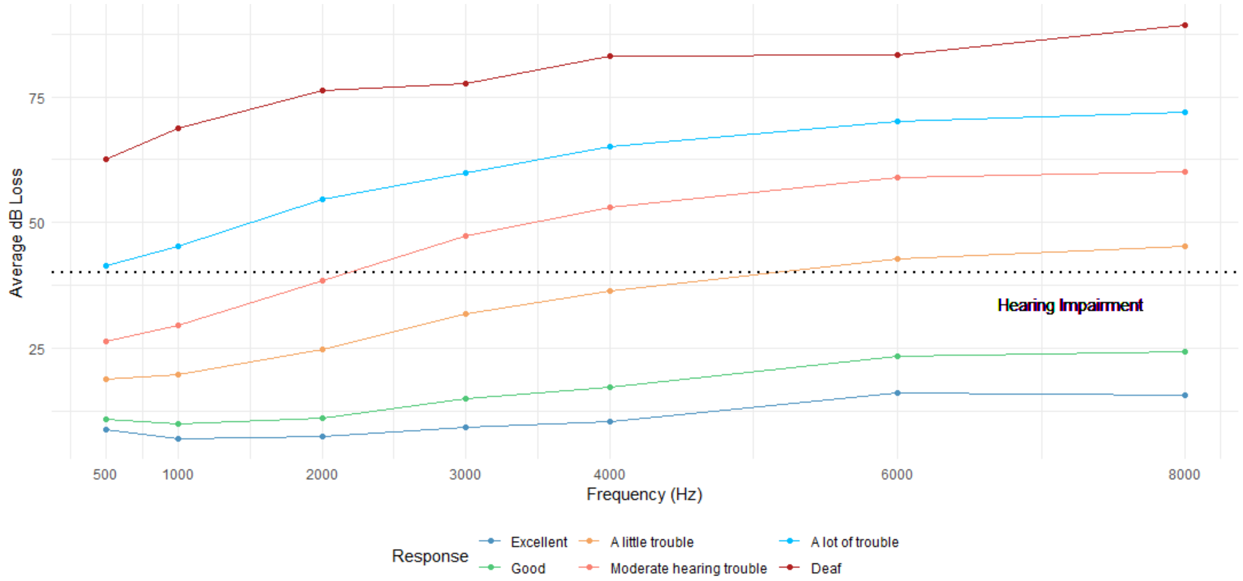


Hearing loss per frequency, stratified by answers. The dotted line represents cut-off value for hearing impairment. Legends: dB: decibel.

**Supplementary Figure 6**

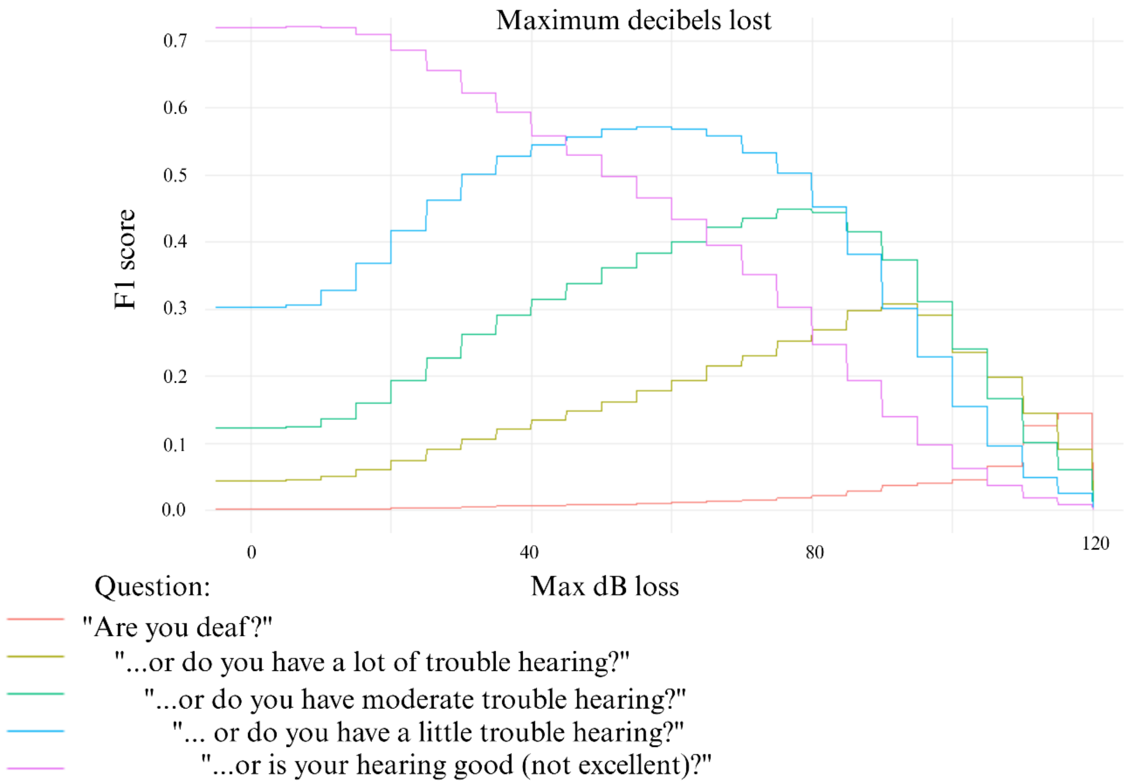


F1 score for different cut-off values of dB loss for common questions.

**Supplementary Figure 7**

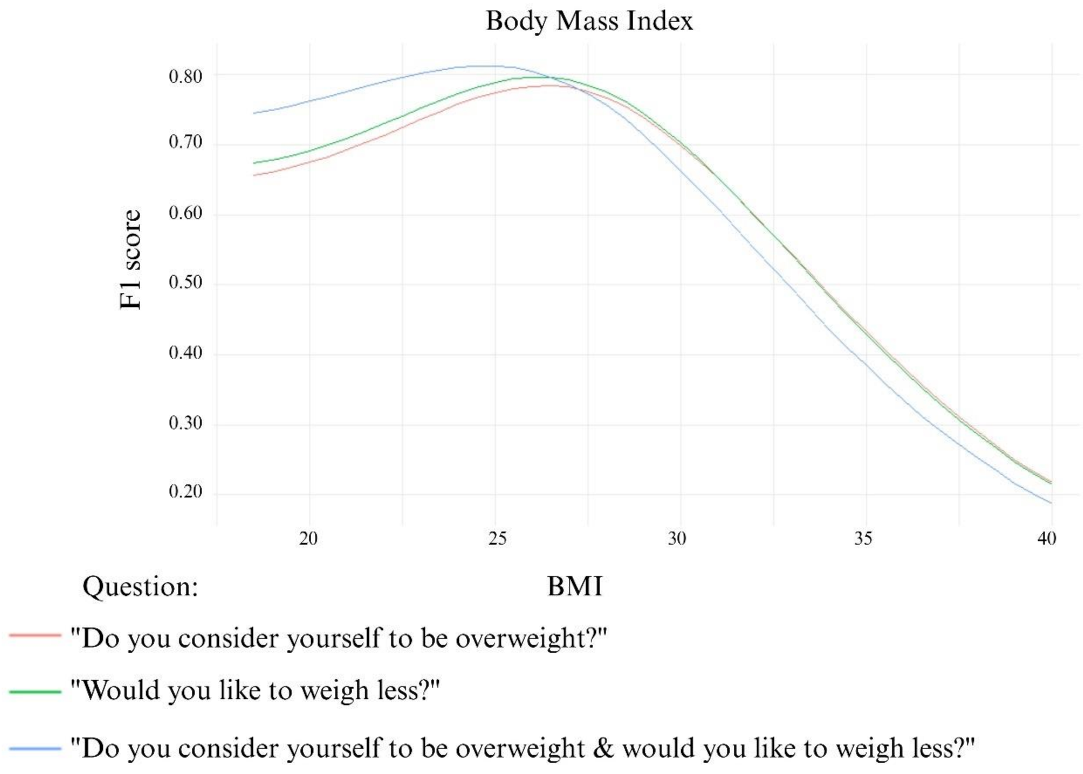


F1 score for different cut-off values of BMI for common questions.
